# An emergency system for monitoring pulse oximetry, peak expiratory flow and body temperature of patients with COVID-19 at home: Development and preliminary application

**DOI:** 10.1101/2020.12.11.20247650

**Authors:** Leonardo Pereira Motta, Pedro Paulo Ferreira da Silva, Bruno Max Borguezan, Jorge Luis Machado do Amaral, Lucimar Gonçalves Milagres, Márcio Neves Bóia, Marcos Rochedo Ferraz, Roberto Mogami, Rodolfo Acatauassú Nunes, Pedro Lopes de Melo

## Abstract

**Background:** COVID-19 is characterized by a rapid change in the patient’s condition, with major changes occurring over a few days. Our aim was to develop and evaluate an emergency system for monitoring patients with COVID-19, which may be useful in hospitals where more severe patients stay in their homes.

**Methodology/Principal findings:** The system consists of the home-based patient unit, which is set up around the patient and the hospital unit, which enables the medical staff to telemonitor the patient’s condition and help to send medical recommendations. The home unit allows the data transmission from the patient to the hospital, which is performed using a cell phone application. The hospital unit includes a virtual instrument developed in LabVIEW^®^ environment that is able to provide a real-time monitoring of the oxygen saturation (SpO_2_), beats per minute (BPM), body temperature (BT) and peak expiratory flow (PEF). Abnormal events may be fast and automatically identified. After the design details are described, the system is validated by a 30-day home monitoring study in 12 controls and 12 patients with COVID-19 presenting asymptomatic to mild disease. Patients presented reduced SpO_2_ (p<0.0001) and increased BPM values (p<0.0001). Three patients (25%) presented PEF values between 50 and 80% of the predicted. Three of the 12 monitored patients presented events of desaturation (SpO_2_<92%). The experimental results were in close agreement with the involved pathophysiology, providing clear evidences that the proposed system can be a useful tool for the remote monitoring of patients with COVID-19.

**Conclusions:** An emergency system for home monitoring of patients with COVID-19 was developed in the current study. The proposed system allowed us to quickly respond to early abnormalities in these patients. This system may contribute to conserve hospital resources for those most in need, while simultaneously enabling early recognition of patients under acute deterioration, requiring urgent assessment.

## 1. Introduction

We are experiencing a global pandemic due to COVID-19 of devastating consequences. The highly infectious pathogen that causes COVID-19, SARS-CoV-2, has infected most of the countries in the world, with over 62.7 million confirmed cases, and just under 1.460.000 deaths as of December 1, 2020 [1].

As the hospital environment becomes more crowded, the criteria for hospital admission become progressively stricter and, as a consequence, more severe patients stay in their homes awaiting improvement or worsening. It was pointed out previously that a rapid clinical deterioration may occur in the initial phase of COVID-19, due to the development of arterial hypoxemia without a concomitant increase in work of breathing [2]. This can prevent an adequate perception by the patient of the real magnitude of the problem. In this context, patients have emerged who silently and rapidly decompensate respiratory function at home, progressing to death even before receiving specialized care. Thus, it is essential to obtain severity markers, especially to predict and prevent the evolution to hospitalization in ICU and death. In this emergency scenario, a consensus has emerged in the literature on the need to institute home monitoring of these patients [3-5], enabling early identification of those who deteriorate acutely and require urgent assessment.

In this context, our aim was to develop an emergency system for monitoring patients with COVID-19, which may be useful in hospitals where more severe patients stay in their homes. We hypothesized that an emergency system based on a smart phone application and specific instruments that allows oxygen saturation, body temperature and peak expiratory flow could be useful as a COVID-19 home-monitoring tool in clinical practice.

## 2. Proposed emergency home monitoring system

Based on previous studies monitoring respiratory diseases [6] and the specific characteristics of COVID-19 [7, 8], the following variables were selected for monitoring:

### 2.1. Pulse Oximetry

In the particular case of COVID-19, this monitoring is essential because in about 10% of cases, especially in the elderly and people with comorbidities, hypoxemia can develop quickly, implying intensive therapy with mechanical ventilation. It is also recognized that hypoxemia is at the very heart of the most severe cases of COVID-19 [9]. In addition, there is now fairly strong evidence that the mortality risk increased with the oxygen saturation reduction [10], which highlights the importance of continuous monitoring of this parameter.

### 2.2. Body temperature

It is known that increased body temperature (BT) is a marker of infection, so temperature provides important orientation of health professionals during the COVID 19 disease course [11]. A recent report by the World Health Organization showed that of the approximately 56,000 laboratory confirmed cases studied in China, among the typical signs and symptoms of COVID-19, increased BT is the most common [12]. Thus, BT is widely recognized in the literature as an important measurement in the COVID-19 pandemic [11-13], that can be used as an easily obtained prognostic indicator [11, 14].

### 2.3. Peak expiratory flow

The monitoring of peak expiratory flow (PEF) is widely recommended in international guidelines for the management of asthma. In these patients, the predicted percentage of PEF correlates reasonably well with the predicted percentage for forced expiratory volume in the first second (FEV1) and provides an objective measure of airflow limitation when spirometry is not available [15]. The value of PFE can be measured using portable meters, which are affordable and relatively simple to handle. This method is widely recognized as suitable for monitoring the progression of the disease and its treatment, using the initial values obtained from the patient as a control [16-18]. The monitoring of PEF also helps to monitor the improvement in the patient after a particular mode of treatment [18]. In general, PEF is reduced in all types of respiratory diseases. PEF has also been a valuable measure in differentiating dyspnea secondary to congestive heart failure versus chronic lung disease [39]. Ignacio-garcia et al. observed a significant improvement in morbidity parameters in the 35 patients with asthma monitored through peak flow at home [19].

In asthma, the drop in PEF values indicates a decrease in the condition of patients. From the initial measurements of PEF, the fall in its value by up to 20% indicates caution, but there is no danger, as this variation is not unexpected in a 24-hour period. A drop of 20 to 50% indicates that the patient is at risk of suffering an exacerbation. If the drop exceeds 50%, patients are at imminent risk of exacerbation. Reduced PEF values in patients with asthma precede the presence of shortness of breath or even the signs of wheezing and snoring detected by the stethoscope. Thus, correct knowledge of PEF predicts the patient’s condition and offers valuable time and opportunity to take all necessary measures to prevent adverse effects of the disease. The integration of PEF monitoring would allow the identification of fall limits of the PEF suitable for monitoring the COVID-19, reducing hospitalizations and death. In this sense, previous studies show that the daily monitoring of PEF was a useful tool in the identification of patients with COPD predisposed to worse clinical outcomes [20].

Previous studies describing the physiological changes in COVID-19 have shown rapid reductions in lung volumes in the presence of edema [8, 21], which highlights the importance of monitoring the PEF of these patients. However, there is no previous studies concerning the use of PEF in COVID-19.

### 2.4. System architecture

The general architecture of the system is reported in Figure 1. The system consists mainly of two parts: 1) the home-based patient unit, which is set up around the patient to acquire data and to receive medical recommendations, and 2) the hospital unit, which enables the medical staff to telemonitor the patient’s condition and help to send medical recommendations.

**Figure 1:**
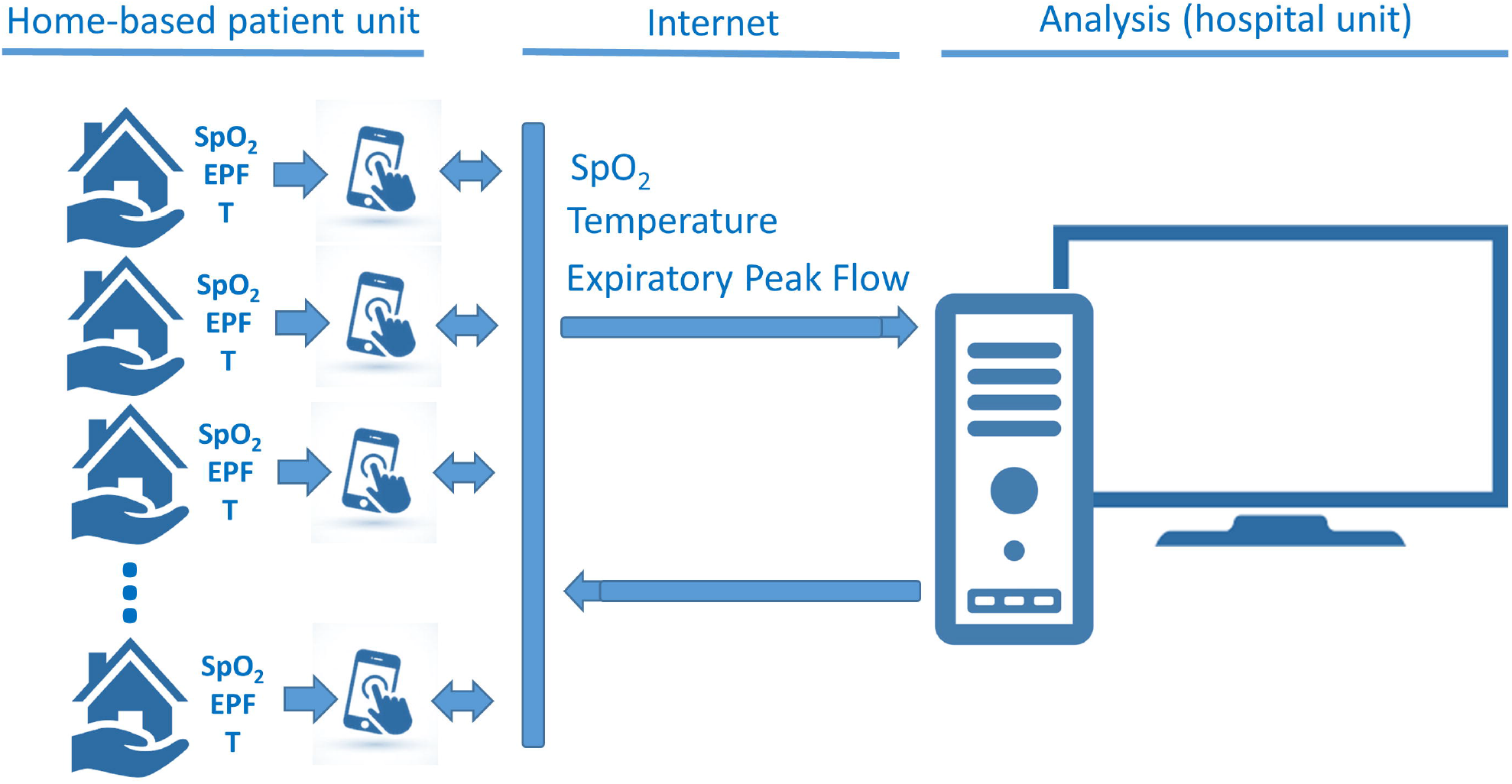
General architecture of the proposed system describing the home-based patient unit, which is set up around the patient to acquire data and the hospital unit, which enables the medical staff to telemonitor the patient’s condition.

Considering the urgency of patient care, the home-based patient unit was developed using readily available commercial instruments. In order to simplify the use of the system by patients, easy-to-use instruments were selected. Thus, patients were assessed using a portable pulse oximeter (finger type, BIC model YK-80A) together with a disposable peak flow meter (Medicate, model 72000M). The used thermometer was the one owned by the patients.

The home unit allows the data transmission from the patient to the hospital, which is performed using a cell phone application. The application was developed in Java using the integrated development environment Android Studio (version 3.6.3). It is based on a form that is filled out and sent by the patient. To make clinical use easier for non-technical personnel, a dedicated user-friendly front panel was developed in the smartphone environment. This interface is shown in Figure 2.

**Figure 2:**
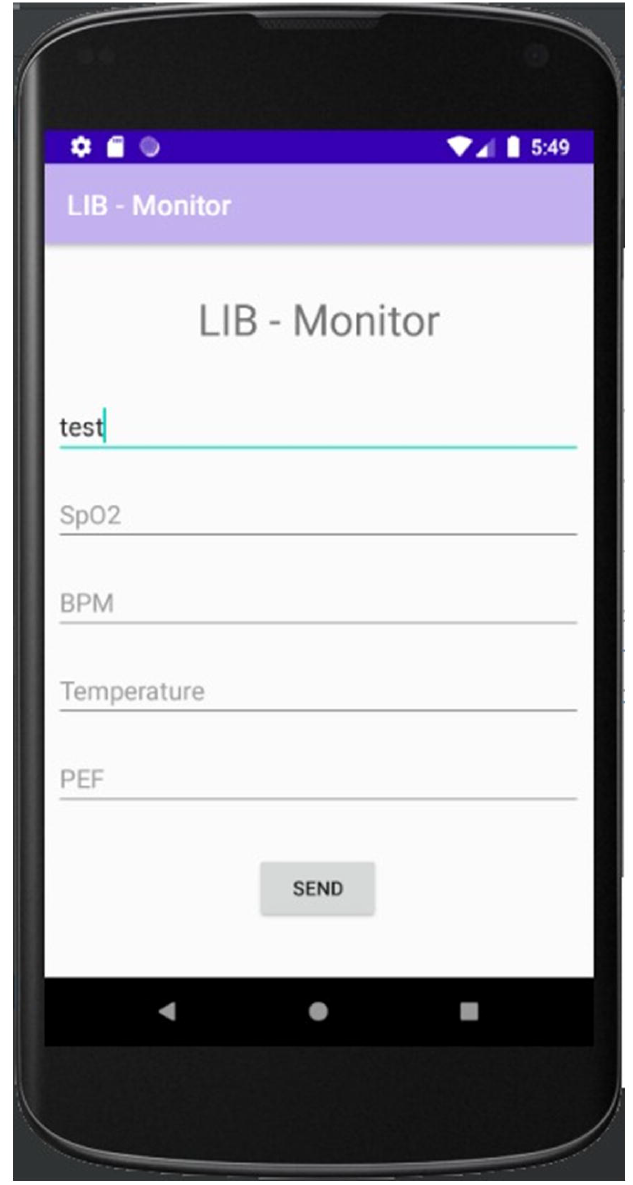
User interface in the cell phone application observed in the home unit. This application allows data transmission from the patient to the hospital. To make clinical use easier for non-technical personnel, a dedicated user-friendly front panel was developed.

After filled out, this form is sent from the application to an online script from Google, the script saves the data in a worksheet according to the patient ID. To this end, the application uses Google Sheets (spreadsheet where data is saved) and Google Scrips (script that integrates the application and Google Spreadsheet). As a result, the application creates an Excel file with one spreadsheet for each patient, which the maximum size is 5 million cells.

Additionally, the values obtained in these exams are also recorded by the patients in a follow-up paper personal diary. These diaries are provided to patients at the beginning of the monitoring. This redundancy is important in order to maintain the perfect functioning of the system even in case of failures in the Internet or other system component.

In the hospital environment, on the other hand, the hardware platform was constituted by an Intel Core i7-8750H, 2.2 GHz computer with 16 GB of RAM, a hard disk of 1 TB and Microsoft Windows 10 operating system. The software was developed in the LABVIEW 2020 environment (National Instruments, Austin, TX). A user-friendly front panel was also developed to be used in the hospital environment. This interface is shown in Figure 3. Its use is described in the flow diagram presented in Figure 4, and the basic LabVIEW program is described in Figure 5.

**Figure 3:**
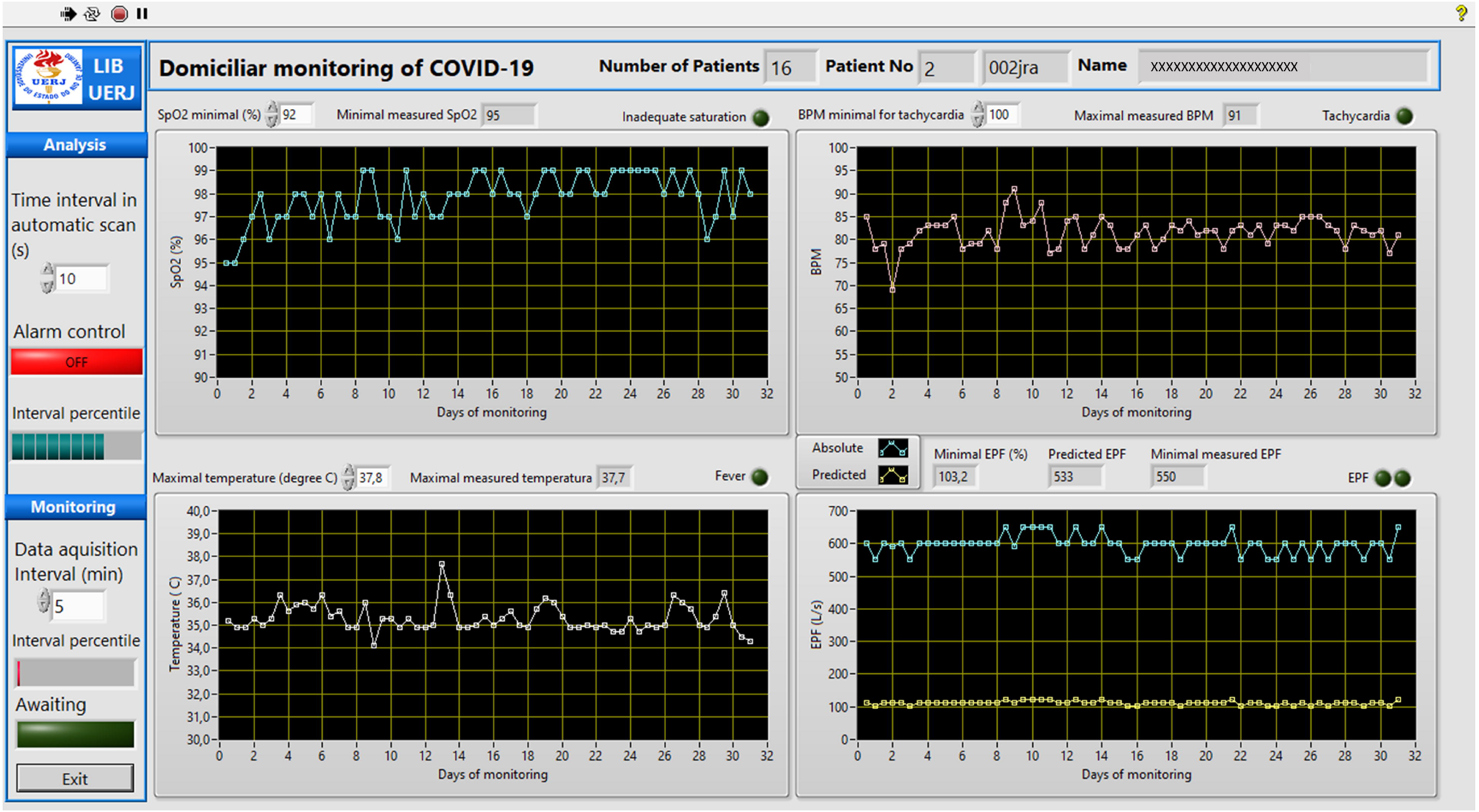
User-friendly front panel that was developed to be used in the hospital environment. The program automatically downloads the patient’s measurements and searches for abnormal values. The limits of abnormality may be adjusted by the user. In the presence of abnormal values, a red indicator light starts flashing, simultaneously with an alarm beep. More detail of this program may be obtained in the flow diagram presented in Figure 4, and the basic LabVIEW program described in Figure 5.

**Figure 4:**
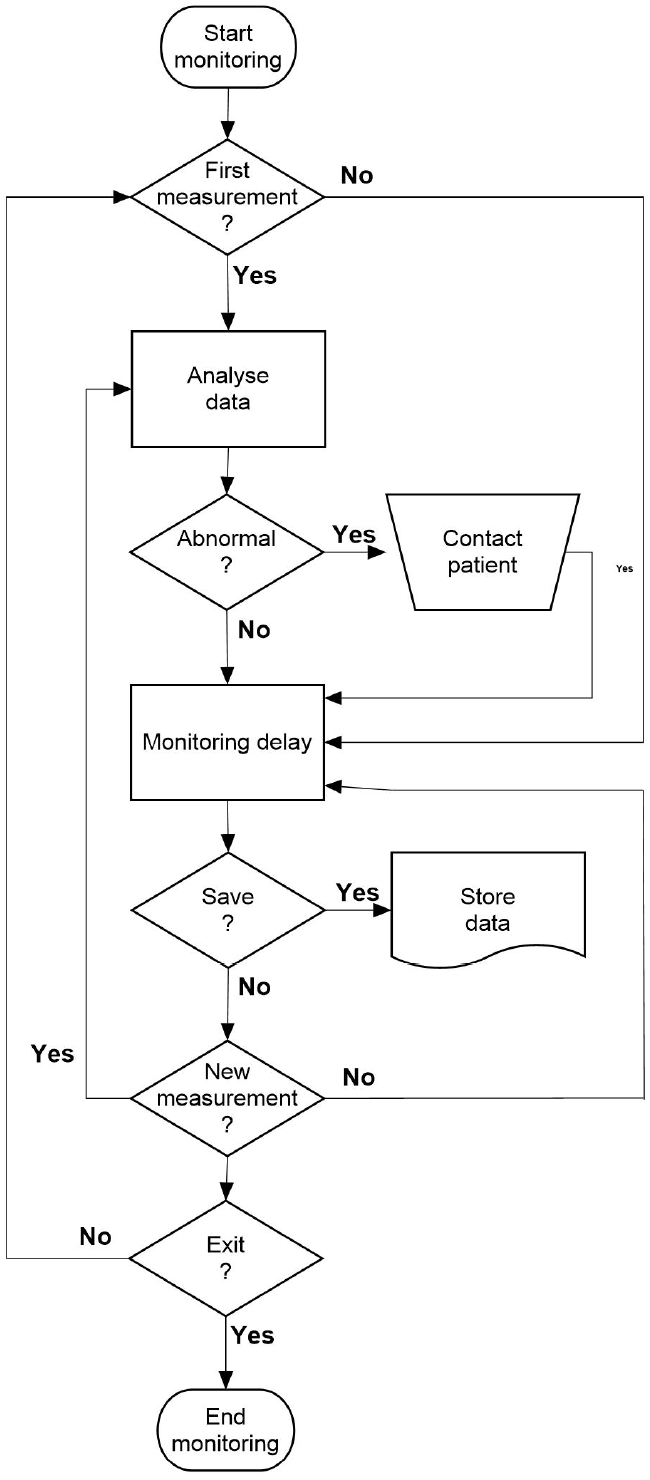
Flow diagram describing the basic operation of the program used in the hospital environment.

**Figure 5:**
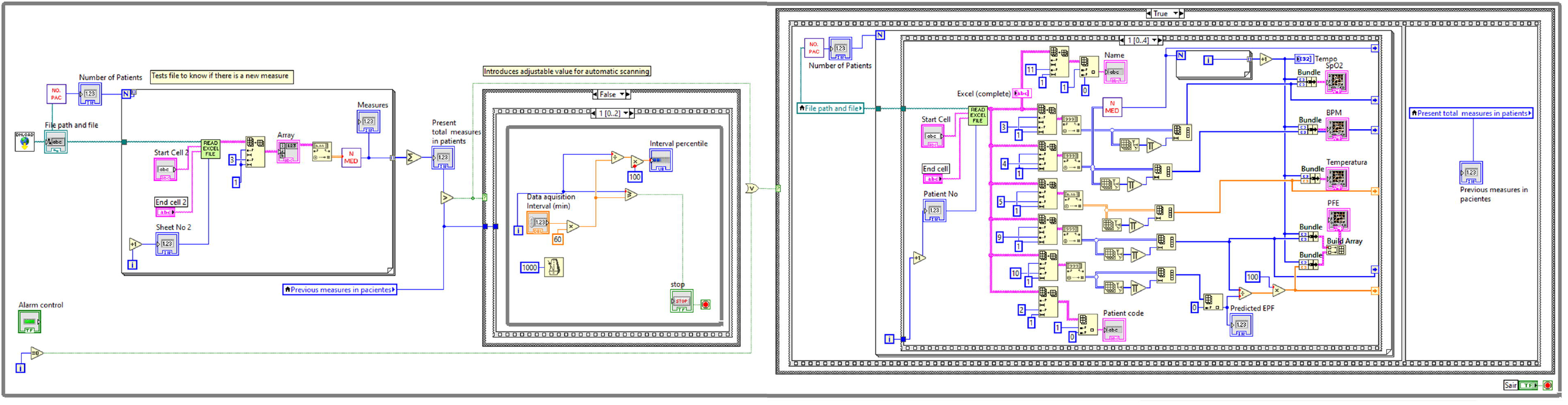
Basic LabVIEW program used in the hospital environment.

The application installed on patients’ cell phones sends the data obtained over the monitoring periods to the analysis center in the hospital, where they are stored. Initially, the program (Figure 5) reads the data of the first patient and elaborates the graphs of SpO_2_, BPM, temperature and PEF (Figure 4). This provides a graphical description of these readings against time (Figure 3), allowing the user to detect trends toward normality or abnormal changes in the mentioned parameters by visual analysis.

The program allows the user to select the minimum SpO_2_ value considered appropriate. Previous studies used a minimum normal SpO_2_ of 92% [22, 23], while others report that these values are between 93% and 94% [10]. The program automatically searches for the minimum SpO_2_ values presented by this patient and compared with the selected value. In the presence of saturation values smaller than the value selected by the user, a red indicator light starts flashing, simultaneously with an alarm beep.

Similar analyses are performed in terms of BPM and temperature. The user can select the maximum BPM value considered appropriate, as well as the maximum temperature considered normal. In this sense, usually values above 100 BPM are indicative of tachycardia [24] and body temperatures above 37 °C are considered abnormal [11, 24]. The system automatically identifies abnormal values presented by this patient and, in the presence of these events, specific red light indicators starts flashing, indicating the nature of the abnormal event, simultaneously with an alarm sound, similar to the indication of abnormal values in SpO2.

EPF analyses are performed taking into account the measured absolute values (blue trace in the EPF chart in Figure 9) and the percent of the predicted values [25] for each monitored patient (yellow trace in the EPF chart in Figure 9). We are performing an initial analysis adapting a methodology traditionally used in asthmatic patients [26]. A zone scheme similar to a traffic light system (green-yellow-red zones) was used to evaluate the predicted EPF values (EPFp) obtained by the patient. The green zone is characterized by PEF readings between 100 to 80% of the EPFp, and signals “all clear”. The yellow zone includes reading from 80 to 50% of the EPFp, and signals “caution”, while the red zone (below 50% of the EPFp) signals “medical alert”. The limits used in the cited zones are probably not adequate for COVID-19 patients, and we hope that they can be rapidly adjusted as the experience of using the system accumulates.

When the system is started and used for the first time on the day, the cited analyses are performed for all patients under home monitoring (Figure 4). To simplify the visual analysis of the results from nontechnical personnel, the software allows the user to perform this first evaluations with an automatic increment among patients and a constant time window for each patient (2 minutes, for example, Figure 3). The automatic increment process can be interrupted if more intriguing results that require a longer analysis time are observed in a given patient. The patient details, monitoring values, and the cited analysis may be saved for further analysis (Figure 4).

After the first scan of patient results, the system automatically updates the results whenever a patient reports new measurements (Figure 4). This is an important feature as it allows the real-time identification of adverse events, and, as a consequence, the clinician may quickly implement the treatment plan. This can be very useful in COVID-19, since this disease is characterized by the presence of rapid deterioration in the patient conditions.

The system also allows the analysis of the results of the measurements in the measurements performed in patients in an asynchronous manner, while monitoring is performed. The medical recommendations, obtained from the analysis of the results sent previously, may be sent using e-mail messages. In case of severely abnormal values, emergency cell phone contact may be used. To help this contact, the system also automatically makes available the patient’s phone number. To receive recommendations in this manner could be a significant benefit for the patient, allowing the fast and easy adjustment of medical treatments. The front panel of this program is similar to that used in the online monitoring system. The interested reader may find a detailed description of this front panel in the supplement (Figure S1).

All the programs used in the current study are available from the corresponding author on request.

## Methods

The Research Ethics Committee of the Pedro Ernesto University Hospital (HUPE) approved the study that obeys the Declaration of Helsinki. The written post-informed consent of all volunteers was obtained before inclusion in the study.

### Monitoring protocol

Based on previous studies on asthma, patients are asked to measure and record their SpO_2_, BPM, body temperature and PEF two times daily, in the morning and in the afternoon, for thirty days. All of the instruments were provided at no cost to the patients.

### Preliminary study

Twenty-four volunteers were selected for the study, including 12 controls and 12 diagnosed with COVID-19. The control group was composed of healthy subjects with no history of COVID-19, tobacco use, as well as cardiac or pulmonary disease. Among COVID-19 patients, all presented asymptomatic to mild disease. All patients were older than 18 years of age, and were enrolled if they had positive COVID-19 testing, which was performed using reverse transcriptase–polymerase chain reaction (RT-PCR) of an oropharyngeal or nasopharyngeal swab. Hospital evaluations after RT-PCR tests showed that all patients had resting SpO_2_ ≥ 94% before the home-monitoring period. In the presence of values indicating adverse events during this period, the need for hospitalization was left to the discretion of the physician evaluating the patient, independent of this study. None of the studied patients required noninvasive or invasive ventilation during or after the monitoring period.

### Statistics

Initially, the sample distribution characteristics were assessed using Shapiro-Wilk’s test. Since data were non-normally distributed, non-parametric analysis (Mann-Whitney test) were performed. Differences with p≤0.05 were considered statistically significant. These analyses were performed using Origin® 8.0 (Microcal Software Inc., Northampton, Massachusetts, United States). Graphs were elaborated using MedCalc 13.1, and the results are present as the median and interquartile range.

### Preliminary results

The biometric characteristics of the studied subjects are described in Table 1, while the past medical history and medication use of the patients with COVID-19 are described in Table 2.

**Table 1:** Mean ± SD of the biometric characteristics of control individuals and patients with COVID-19.

|  | Age<br>(years) | Body mass<br>(kg) | Height<br>(cm) | BMI<br>(kg/m<sup>2</sup>) | Gender<br>(M/F) |
| --- | --- | --- | --- | --- | --- |
| Control (n=12) | 38.2±15.0 | 68.2±10.7 | 167.8±7.2 | 24.2±3.1 | 4/8 |
| COVID-19 (n= 12) | 37.2±13.3 | 77.6±16.4 | 163.8±8.2 | 28.9±5.4 | 3/9 |
| p | ns | ns | ns | 0.02 | - |

**Table 2:** Characteristics of COVID-19 patients.

|  | N | (%) |
| --- | --- | --- |
| Smoking | 1 | 8.3 |
| Overweight | 7 | 58.3 |
| Obesity | 3 | 25.0 |
| Controlled diabetes | 1 | 8.3 |
| Pre-diabetes | 1 | 8.3 |
| Hypertension | 2 | 16.7 |
| Epilepsy | 1 | 8.3 |
| Cardiac arrhythmias | 1 | 8.3 |
| Essential thrombocytosis | 1 | 8.3 |
| Received hydroxychloroquine and azithromycin | 5 | 41.2 |
| Ivermectin | 3 | 25.0 |

Overweight was defined as body mass index (BMI) between 25 and 30 kg/m<sup>2</sup>; Obesity was defined as BMI ≥ 30 kg/m<sup>2</sup>; N, number of patients; %: percentage of the total number of patients.

Figures 6 and 7 describes the results of the SpO_2_ and BPM in controls and patients, respectively.

**Figure 6:**
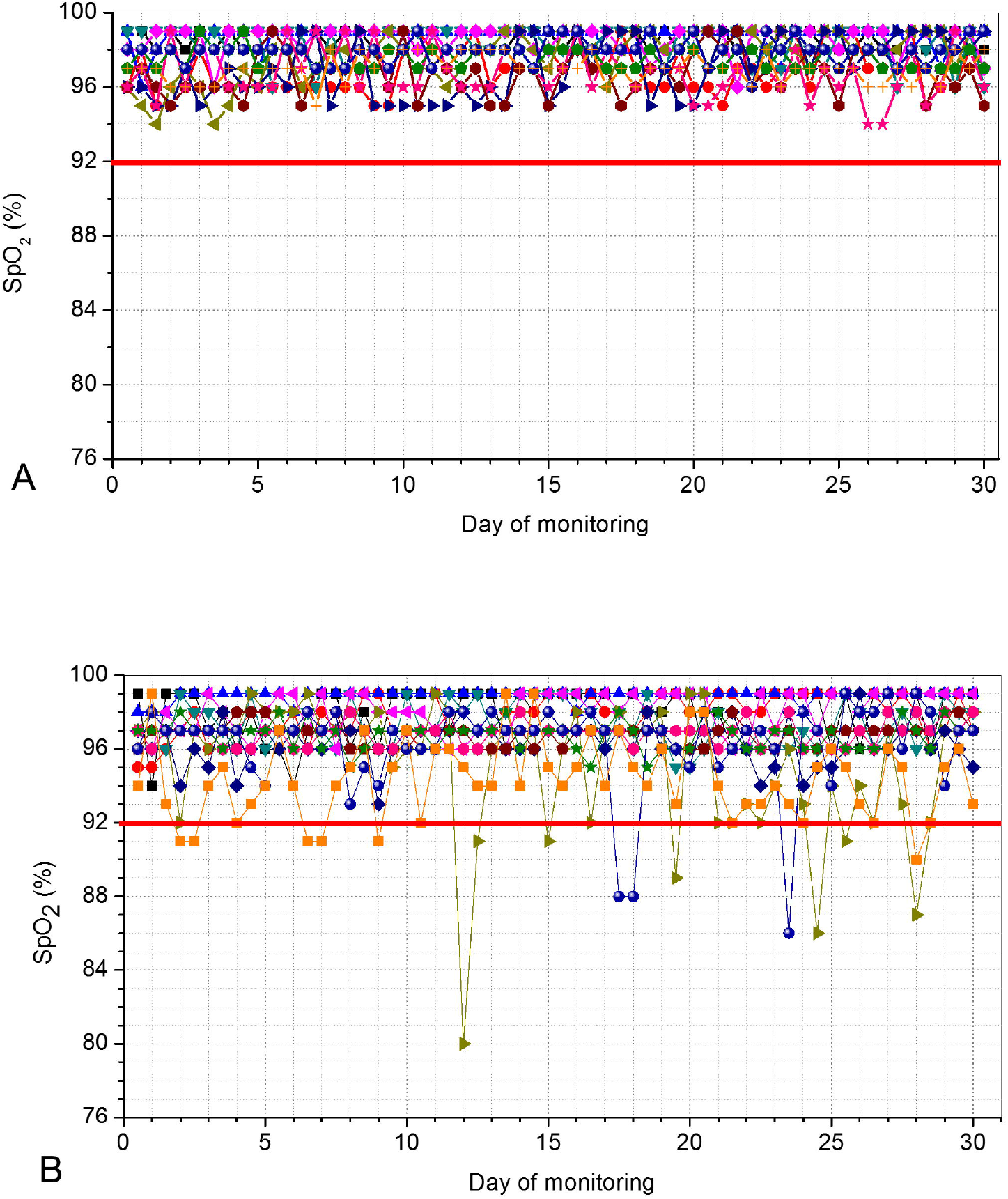
Home pulse oximetry values telemonitored during a 30-days period in controls (A) and in COVID-19–positive patients. (B). The red line describes the limit considered as a minimum normal value of saturation (92%). Three patients presented values bellow this limit.

**Figure 7:**
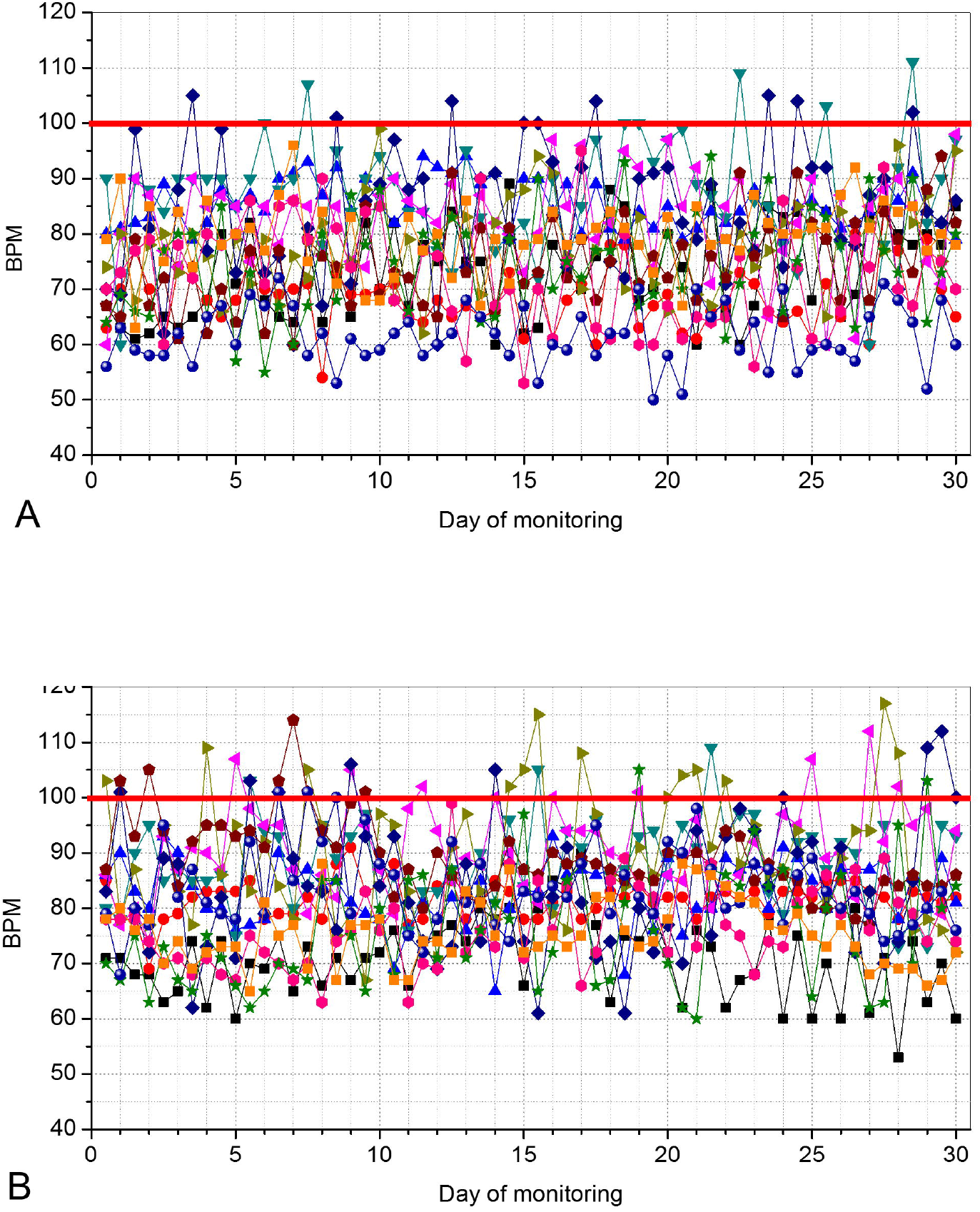
Beats per minute (BPM) values telemonitored during a 30-days period in controls (A) and in COVID-19–positive patients (B). The red line describes the limit considered as a maximum normal value of BPM (100 BPM). Seven patients presented values above this limit along this home readings.

COVID-19 resulted in a significant reduction in SpO_2_ (Figure 8A, p<0.0001) and increased values of BPM (Figure 8B, p<0.0001).

**Figure 8:**
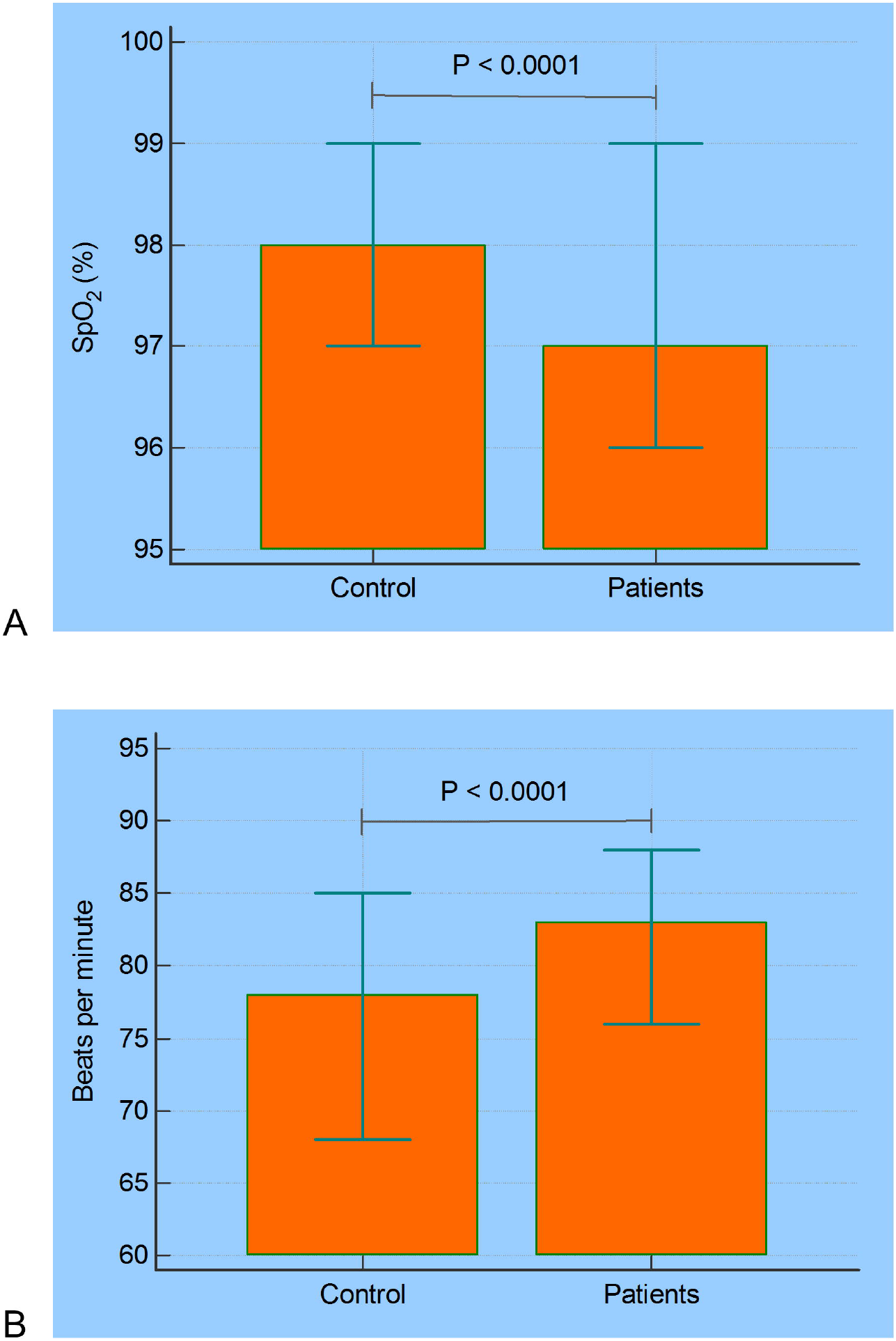
Median and 95% confidence intervals of the home monitored oxygen saturation (A) and BPM (B) in controls and COVID-19–positive patients.

**Figure 9:**
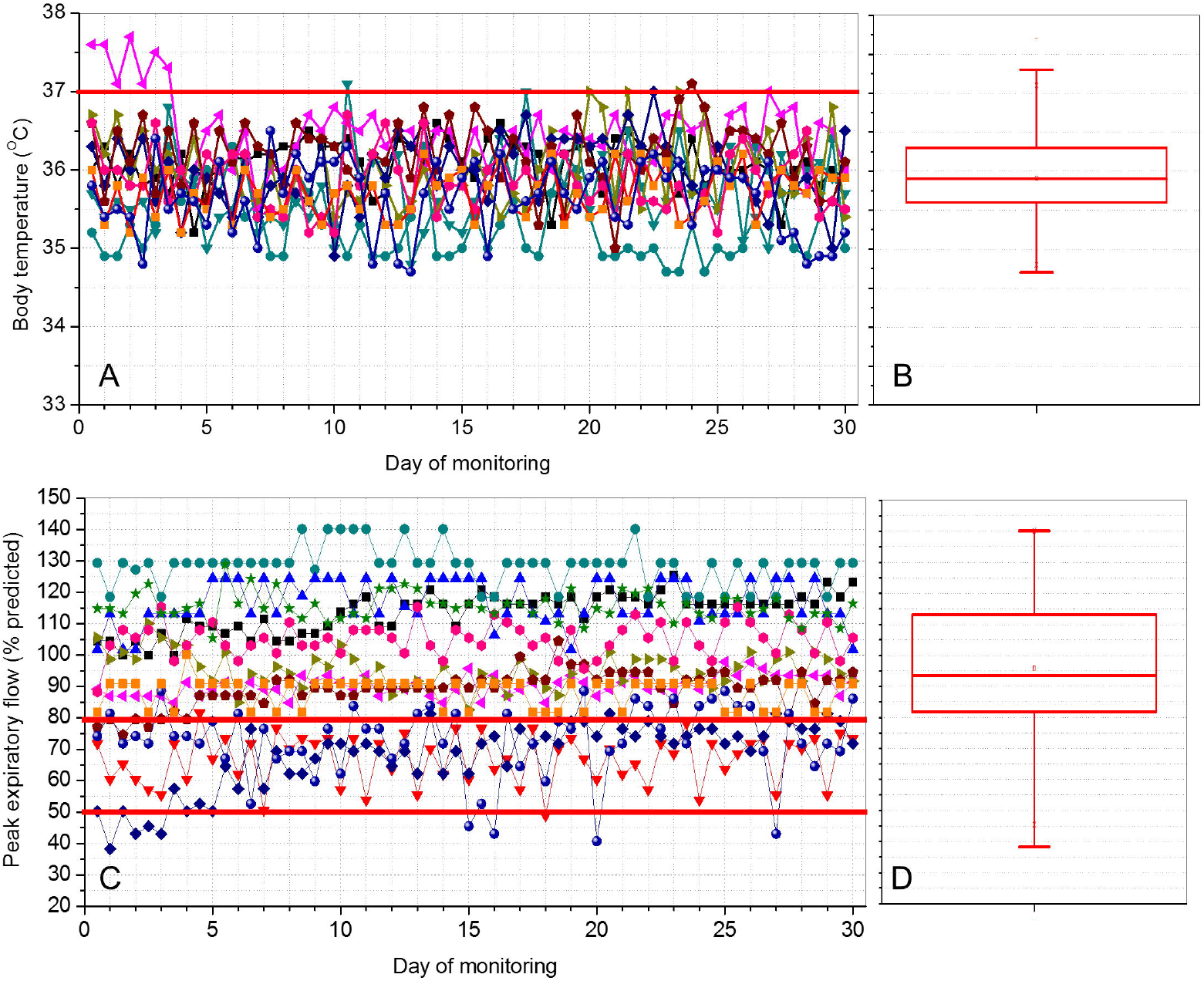
Results obtained in the daily telemonitoring of temperature (A) and peak expiratory flow (C) in COVID-19–positive patients during 30 days. Box-plot descriptions of these results are also presented for the temperature (B) and peak expiratory flow (D). The top and the bottom of the box plot represent the 25th-to 75th-percentile values while the circle represents the mean value, and the bar across the box represents the 50th-percentile value.

Figure 9 shows the results obtained in the daily monitoring of temperature and PEF in patients with COVID-19 during 30 days. Most of the studied patients presented PEF values above 80% during the monitoring period (8 patients, 66.7%), while 3 patients (25%) had values between 50 and 80% of predicted (Figure 9). Two of the 12 patients (17%) showed transient values below 50%, which subsequently evolved to values between 50 and 80% of the predicted values.

## Discussion

To the best of our knowledge, this is the first study to provide a detailed description of an emergency remote monitoring system for individuals with COVID-19. The system is able to provide a real-time monitoring of oximetry, BPM, body temperature and peak expiratory flow. The preliminary results obtained in the 30-day validation study showed that patients with COVID-19 presented reduced SpO_2_ and PEF values, as well as increased BPM values. This was also the first study to investigate the use of PEF in COVID-19. The proposed system allowed us to quickly respond to early abnormalities in patients with COVID-19.

During the 30-days period of the initial tests of the proposed system, 720 data-points regarding SpO_2_ were remotely obtained, resulting in a total of 16 alerts among the 12 monitored patients (Figure 6). It was observed that these alerts resulted from an abrupt drop in SpO_2_ rather than a gradual decline. This is consistent with previous results [23], and is probably associated with the rapid deterioration caused by a surge in proinflammatory molecules in the “cytokine storm” phase of COVID-19 [27].

The values of SpO_2_ were reduced in patients with COVID-19 in comparison with controls (Figure 8A). This finding is consistent with the observation that in the initial phase of COVID-19 there is an increase in V/Q mismatch and thus persistence of pulmonary arterial blood flow to non-ventilated alveoli. The current understanding is that this results from the infection, which leads to a modest local interstitial edema and loss of surfactant. These factors are associated with alveolar collapse and intrapulmonary shunting [2]. The results observed in Figure 6 are in line with that obtained by O’Carroll and collaborators [28] investigating the remote monitoring of SpO_2_ in individuals with COVID-19.

It was pointed out previously that data are lacking for young adults who often present with mild or asymptomatic disease, a part of the population considered to be highly contagious [29]. Figures 7 and 8B contributes to elucidate this question providing evidence that COVID-19 introduces increased values of BPM in comparison with control subjects (Figure 8B). These results are consistent with a previous review showing that COVID-19 is related with a number of cardiovascular complications [30], and the use of increased heart rate as a clinical criteria for hospital admission in COVID-19 pneumonia [31].

It is known that the degree of temperature elevation might reflect the severity of inflammation [11]. Previous studies suggest that poor BT control during the COVID 19 disease course is a marker of poor prognosis and BT can be used as an easily obtained prognostic indicator [11]. At this point in the COVID-19 outbreak, however, a specific fever pattern associated with this disease has not yet been identified. In fact, there is a paucity of data on temperature management for COVID-19 [14]. The results of the present study (Figures 9A and B) indicate that the proposed system can help to fill this important gap in the literature. Low fever events were observed in 5 of the 12 studied patients (41.7%), which is consistent with the characteristics of asymptomatic to mild disease of the studied COVID-19 group.

Figure 9C and D shows that most of the studied patients presented normal PEF values during the monitoring period. Patients who have been infected by the Coronavirus may develop pulmonary edema and atelectasis, which my result in a reduction in lung volume. Thus, it may be speculated that this reduction may introduce a limitation of the expiratory flow influencing the values obtained in peak-flow measurements. This processes would be similar to that observed in restrictive diseases in which the volumes exhaled are reduced [32]. These effects may explain, at least in part, the decrease in PEF presented in three of the studied patients, as well as the transient values below 50% of the predicted values observed in two patients, as described in Figure 9A. It is noteworthy that one of the patients who had EPF <50% also had desaturation <92%.

There is general agreement in the literature that, given the severity of the ongoing global pandemic, the ability to remotely monitor patients who do not require hospitalization is essential for optimal utilization of health care resources [23]. To contribute in this direction, this study presents a low-cost open-architecture emergency system for remote monitoring of patients with COVID-19. There are currently no data to guide the use of home pulse oximetry in COVID-19 patients or its validity in identifying disease progression [23]. There is also a paucity of data on temperature management for COVID-19. No previous study has investigated the use of EFP in COVID-19. The system presented in this study may help to quickly accumulate data on SpO_2_, body temperature and EPF, contributing to the development of the guidelines for these clinical practices.

Remote monitoring systems has been increasingly used in respiratory diseases [6, 33-36]. An important factor, in the particular case of COVID-19, is that the disease is characterized by a rapid change in the patient’s condition, with major changes occurring over a few days. The proposed system may provide an early detection of patient’s conditions deteriorating at home. The use of this technology allowed us to facilitate discharge in patients with mild-to-moderate disease in a safe and appropriate manner. This procedure hold the potential to increase bed availability without compromising safe patient care.

We acknowledge the limitations of our study, including unknown methods of temperature measurement. Nevertheless, a clear trend in increased mortality among the patients with poor temperature control highlights the usefulness of this noninvasively and easily obtained parameter for evaluating patients’ prognoses.

Secondly, one could argue that the study presents a small sample size, and additional studies, including a more significant number of subjects are necessary. These studies would allowed us to perform a detailed investigation concerning the utilization of home pulse oximetry, body temperature and peak expiratory flow monitoring to identify robust predictors of hospitalization.

Finally, the system validation was performed in subjects from a Brazilian population at a single practice site, which affects the study’s generalizability. Therefore, multicenter studies are necessary in the future to expand the generalizability of these findings. The study used broad inclusion criteria and was performed in a typical setting under usual clinical procedures, which enhanced its generalizability.

## Conclusion

An emergency system for home monitoring of SpO_2_, body temperature and PEF in patients with COVID-19 was developed in the current study. This was the first study to propose such a system and to evaluate the use of PEF in COVID-19. Using this system, the acquisition and analysis of the cited signals can be performed remotely through the Internet. The ability of the system to detect abnormal events was initially validated by a 30-day monitoring study in normal subjects and patients with COVID-19. In close agreement with previous results and physiological fundamentals, the presence of COVID-19 resulted in reduced values of SpO_2_, increased BPM, fever events in 41.7% of the patients and decreased PEF in 33% of the studied patients.

The proposed system may contribute to conserve hospital resources for those most in need, while simultaneously enabling early recognition of patients under acute deterioration, requiring urgent assessment.

Based on these promising results, future work includes a clinical trial in which we will perform a follow up in well-defined groups of patients with COVID-19. This will provide a detailed evaluation of the clinical contribution of the home monitoring approach in improving the patient’s care and outcomes.

## Data Availability

The data will be available from a public repository (Open Science Framework) after acceptance.

---

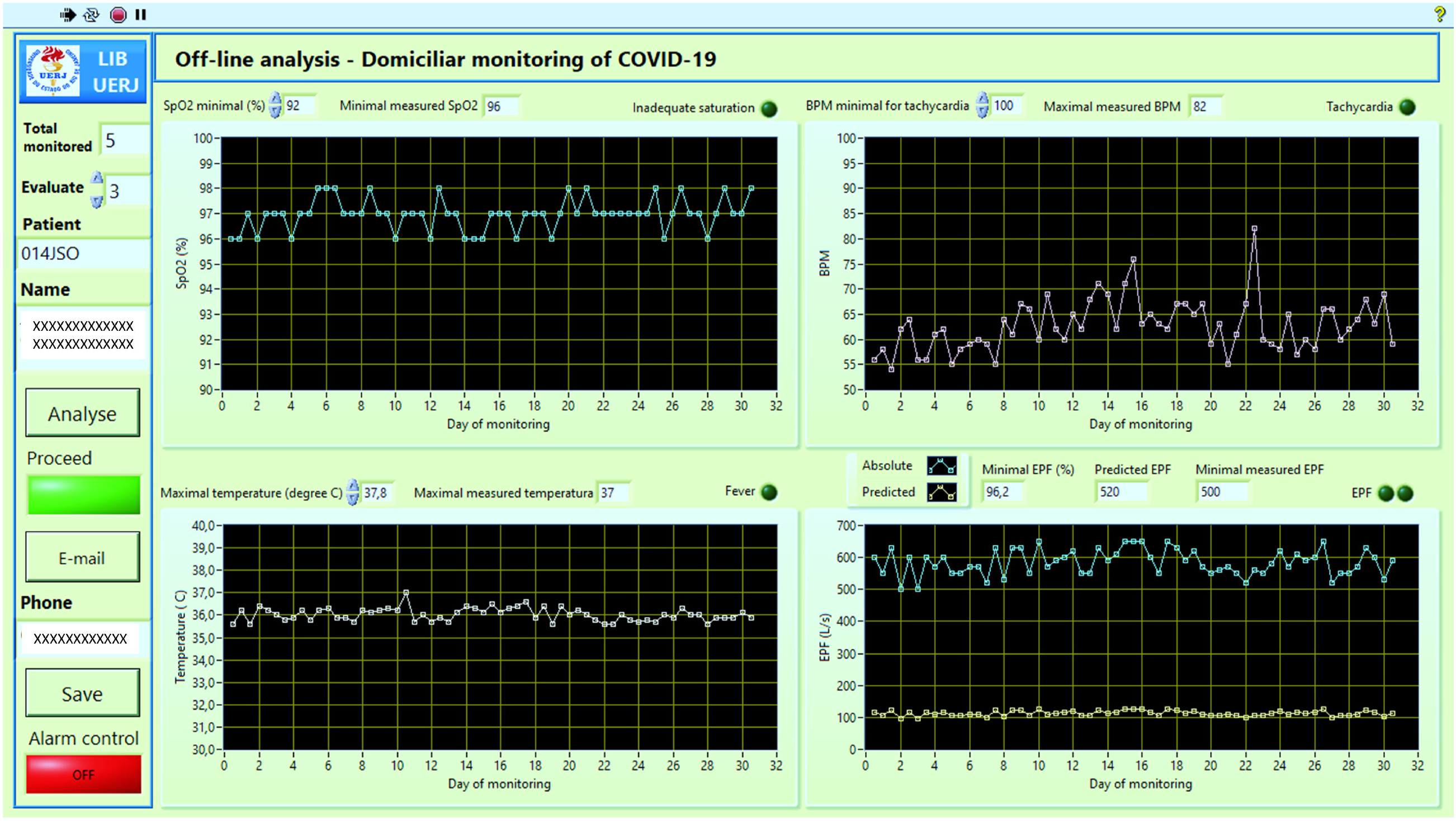

